# Leveraging open tools to realize the potential of self-archiving: A cohort study in clinical trials

**DOI:** 10.1101/2023.01.03.23284111

**Authors:** Delwen L. Franzen

**Affiliations:** Berlin Institute of Health at Charité – Universitätsmedizin Berlin, QUEST Center, Charitéplatz 1, 10117 Berlin, Germany

**Keywords:** green open access, self-archiving, clinical trial, scholarly communication, shareyourpaper, unpaywall

## Abstract

While Open Access (OA) is growing, many publications remain behind a paywall. This limits the impact of research and entrenches global inequalities by restricting access to knowledge to those that can afford it. Many journal policies allow researchers to make a version of their publication openly accessible through self-archiving in a repository, sometimes after an embargo period (green OA). Unpaywall and Shareyourpaper are open tools that help users find OA articles and support authors to legally self-archive their papers, respectively. This study leveraged these tools to assess the potential of green OA to increase discoverability in a cohort of clinical trial results publications from German university medical centers. Of the 1,897 publications in this cohort, 46% (n=871/1,897, 95% Confidence Interval (CI) 44% to 48%) were not openly accessible via a journal or a repository. Of these, 85% (n=736/871, 95% CI 82% to 87%) had a permission to self-archive the accepted or published version in an institutional repository. Thus, most of the closed-access clinical trial results in this cohort could be made openly accessible, in line with World Health Organization (WHO) recommendations. In addition to providing further evidence of the unrealized potential of green OA, this study demonstrates the use of open tools to obtain actionable information on self-archiving at scale, and empowers efforts to increase science discoverability.

## 1. Introduction

Open Access (OA) refers to the free online access, and largely unrestricted sharing, and re-use of scholarly research [1]. While there is evidence that OA is growing [2–6], many publications remain hidden behind a paywall in subscription journals. This limits the reach and impact of research, and entrenches global inequalities by restricting access to knowledge to institutions and individuals that can afford it [7]. The UNESCO Recommendation on Open Science adopted in 2021 outlined several priority areas of action towards achieving open science globally. This included the recommendation to support non-commercial publishing models and promote existing flexibilities in intellectual property systems to broaden access to knowledge for the benefit of scientists and society [8].

One way of increasing access to published research is self-archiving in an OA location (green OA) [9]. In many cases, publisher or journal self-archiving policies allow researchers to make a version of their publication openly accessible, sometimes after an embargo period. These policies typically outline several permissions that differ as to *what* version can be archived, *where* it can be archived, and *when* it can be archived. Self-archiving is also enabled through national or consortia-based licensing of electronic journals [10] as well as through author rights-retention, with some universities having adopted rights-retention OA policies that make it unnecessary to obtain permission from publishers [11]. Moreover, several countries have introduced clauses in copyright law that allow researchers to make a version of their publication openly available under certain conditions, regardless of publisher policies [12,13].

The potential of self-archiving to broaden access to research has been clearly demonstrated. An analysis of self-archiving policies of the largest 100 publishers by output volume found that 80.4% of 1.1 million articles published in subscription-based journals could be shared as a postprint (peer-reviewed version, either accepted or published) in a repository one year after publication [14]. At the same time, a synthesis of previous studies estimated realized green OA to be around 12% [9], suggesting a largely unrealized potential of green OA. Factors contributing to the limited uptake of self-archiving are thought to include a lack of awareness [15] and concerns over copyright infringement [16]. Embargoes on self-archiving specific versions may also contribute to limited uptake of this practice. Suggestions to increase self-archiving have included introducing funder and institutional mandates [17] and providing tools and services that make it quick and easy to accomplish [18].

Efforts to assess and bridge the gap between opportunity and practice have been supported by the development of resources such as SHERPA/RoMEO, which aggregates self-archiving policies at the level of journals in a machine-readable format. These resources are increasingly being integrated with libraries’ deposit systems, albeit in different ways [10,19–21]. Building on these resources, Shareyourpaper (https://shareyourpaper.org, OA.Works) is a tool that supports authors to legally self-archive their papers by distilling applicable policies into machine-readable self-archiving permissions at the level of individual articles. If a permission is found, the paper is automatically deposited in the generalist repository Zenodo. All authors need to do is provide the requested version of the paper. While Shareyourpaper can be integrated with institutional repositories and is therefore scalable for libraries, the deposit in Zenodo also empowers efforts to increase self-archiving beyond a given institution (see more information at [22]). Taken together, Shareyourpaper’s approach of bringing together automated permissions checking and an automated deposit workflow makes it a promising tool to increase self-archiving at scale.

In this study, Shareyourpaper was used in combination with Unpaywall (OurResearch), one of the most established mechanisms to determine the OA status of publications, to assess the potential of self-archiving to increase the discoverability of clinical trial results. Clinical trials are the backbone of evidence-based medicine. They inform regulators, public health agencies, and doctors of which interventions are safe and effective to use. Public health crises have repeatedly highlighted the practical and ethical importance of providing equitable access to the outputs of health and clinical research [23,24]. The World Health Organization (WHO) Joint Statement on Public Disclosure of Results from Clinical Trials states that “[…] publications describing clinical trial results should be open access from the date of publication, wherever possible” [25]. This raises the following research questions:

– **RQ1:** How many clinical trial results publications are openly accessible?
– **RQ2:** For clinical trial results publications that are closed-access, what is the potential of self-archiving to achieve compliance with WHO guidelines?

These research questions were addressed building on a validated cohort of publications describing the results of clinical trials conducted at German university medical centers [26]. This study focused on the potential of self-archiving based on journal and publisher policies. While the right of secondary publication introduced in German copyright law is in principle a powerful tool to drive self-archiving at scale, its use in practice has been limited [27] and implementation attempts have generated as yet unresolved legal disputes [28,29]. Furthermore, self-archiving mandates at the level of German research institutions and funders remain uncommon. In this context, taking advantage of journal and/or publisher self-archiving policies remains an important avenue to broaden access to past and future research.

## 2. Materials and Methods

### 2.1. Trial and publication screening

This study used the previously developed IntoValue cohort of clinical trials and associated publications [26]. This cohort comprises interventional clinical trials registered in ClinicalTrials.gov or the German Clinical Trials Register (DRKS), conducted at a German university medical center, and reported as complete on the registry between 2009 – 2017. In line with WHO definitions, trials in this cohort include all interventional studies and are not limited to highly regulated drug trials [30]. The earliest results publications associated with these trials were found through manual searches [31,32]. As updated registry data was downloaded on 1 November 2022, the IntoValue inclusion criteria were re-applied: interventional, study completion date between 2009 and 2017, complete based on study status, and conducted by a German university medical center. The sample was further limited to journal publications with a unique Digital Object Identifier (DOI) that resolved in Unpaywall and were published between 2010 and 2020.

### 2.2. Determination of OA status

The OA status of publications in this cohort was obtained with Unpaywall. Unpaywall harvests content from legal sources such as publishers, repositories, and preprint servers, and has limited coverage of personal websites. It does not harvest content from academic social networks for which concerns have been raised about the persistence of content [2,33]. Thus, the following definition of OA was used in this paper: articles that are free to read online in a journal or OA repository. Unpaywall was queried via its API using the R package, UnpaywallR (https://github.com/quest-bih/unpaywallR), and all available OA locations were extracted for each publication. These include gold (openly available in an OA journal), hybrid (openly available under an open license in a subscription-based journal), green (openly available in a repository), bronze (openly available on the journal page but without a clear open license), and closed-access. As publications can have several OA types, a hierarchy was applied such that only one OA type was assigned to each publication, in descending order: gold, hybrid, bronze, green, and closed-access. Thus, green OA in this study refers to publications that are only openly accessible via a repository. **Table A1** outlines how OA types were derived in this study. Unpaywall was queried via its API on 17 December 2022.

### 2.3. Determination of the potential to self-archive

To obtain self-archiving permissions for the publications in the cohort, Shareyourpaper was queried via its API using a custom-made Python script (https://github.com/delwen/oa-archiving-permissions). The ‘best permission’ in the API response focuses on how a paper can be self-archived in an institutional repository. Here, publications were defined to have the potential for green OA if a ‘best permission’ was found for archiving either the accepted or published version in an institutional repository, and if the embargo (if applicable) had elapsed by the query date. Permissions for the submitted version were not considered. Publications without a ‘best permission’ and publications with a ‘best permission’ but no information on the embargo period, archiving location, or version were considered as unclear. **Table 1** outlines how article-level permissions were derived in this study. Shareyourpaper was queried via its API on 17 December 2022.

**Table 1.** Criteria used to derive self-archiving permissions based on the Shareyourpaper API query.

| Permission | Combination of fields |
| --- | --- |
| Permission to archive the accepted version in an institutional repository (based on “best permission” | `can_archive = TRUE AND “institutional repository” IS IN locations AND (embargo_months = 0 OR embargo has elapsed) AND “acceptedVersion” IS IN versions` |
| Permission to archive the published version in an institutional repository (based on “best permission”) | `can_archive = TRUE AND “institutional repository” IS IN locations AND (embargo_months = 0 OR embargo has elapsed) AND “publishedVersion” IS IN versions` |

The realized potential of green OA was estimated based on a) the number of publications that were only openly accessible in a repository (green OA), and b) the number of closed-access publications for which a self-archiving permission was found (based on the criteria defined in **Table 1**). By virtue of being archived, green OA publications were assumed to have had a permission for self-archiving in a repository. Neither the version nor the OA location of green OA publications was systematically checked.

### 2.4. Software, code, and data

Data processing was performed in R (version 4.0.5) [34] and Python 3.9 (Python Software Foundation, Wilmington, DE, USA). All the code generated in this study is available in GitHub under an open license (https://github.com/delwen/oa-archiving-permissions). The data presented in this study are openly available in Zenodo [35].

## 3. Results

### 3.1. Trial and publication screening

The IntoValue dataset includes interventional trials registered in ClinicalTrials.gov or DRKS, conducted at a German university medical center, and completed between 2009 and 2017 (n = 3,788). After applying the exclusion criteria, the sample included 1,897 unique trial publications that resolved in Unpaywall and were published between 2010 and 2020. **Figures B1-B2** provide flow diagrams of the trial and publication screening, respectively.

### 3.2. OA status of publications

Of the 1,897 clinical trial results publications examined in this study, 54% (n = 1,026/1,897, 95% CI 52% to 56%) were openly accessible in a journal (gold, hybrid, bronze) or in a repository (green). Across all years, the cohort included 432 gold OA (23%, 95% CI 21% to 25%), 141 hybrid OA (7%, 95% CI 6% to 9%), 310 bronze OA (16%, 95% CI 15% to 18%), and 143 green OA (8%, 95% CI 6% to 9%) publications (**Figure 1**). The smaller contribution of green OA likely reflects the hierarchy of OA locations used, whereby articles that were openly accessible both in a journal and in a repository were not counted as green OA. The proportion of openly accessible publications increased from 50% (n = 19/38, 95% CI 35% to 65%) in 2010 to 73% (n = 67/92, 95% CI 62% to 81%) in 2020. This largely appeared to be due to an increase in gold and hybrid OA. In turn, across all publication years, 46% (n = 871/1,897, 95% CI 44% to 48%) of publications were not openly accessible via a journal or repository (**Figure 1**) (RQ1).

**Figure 1.**
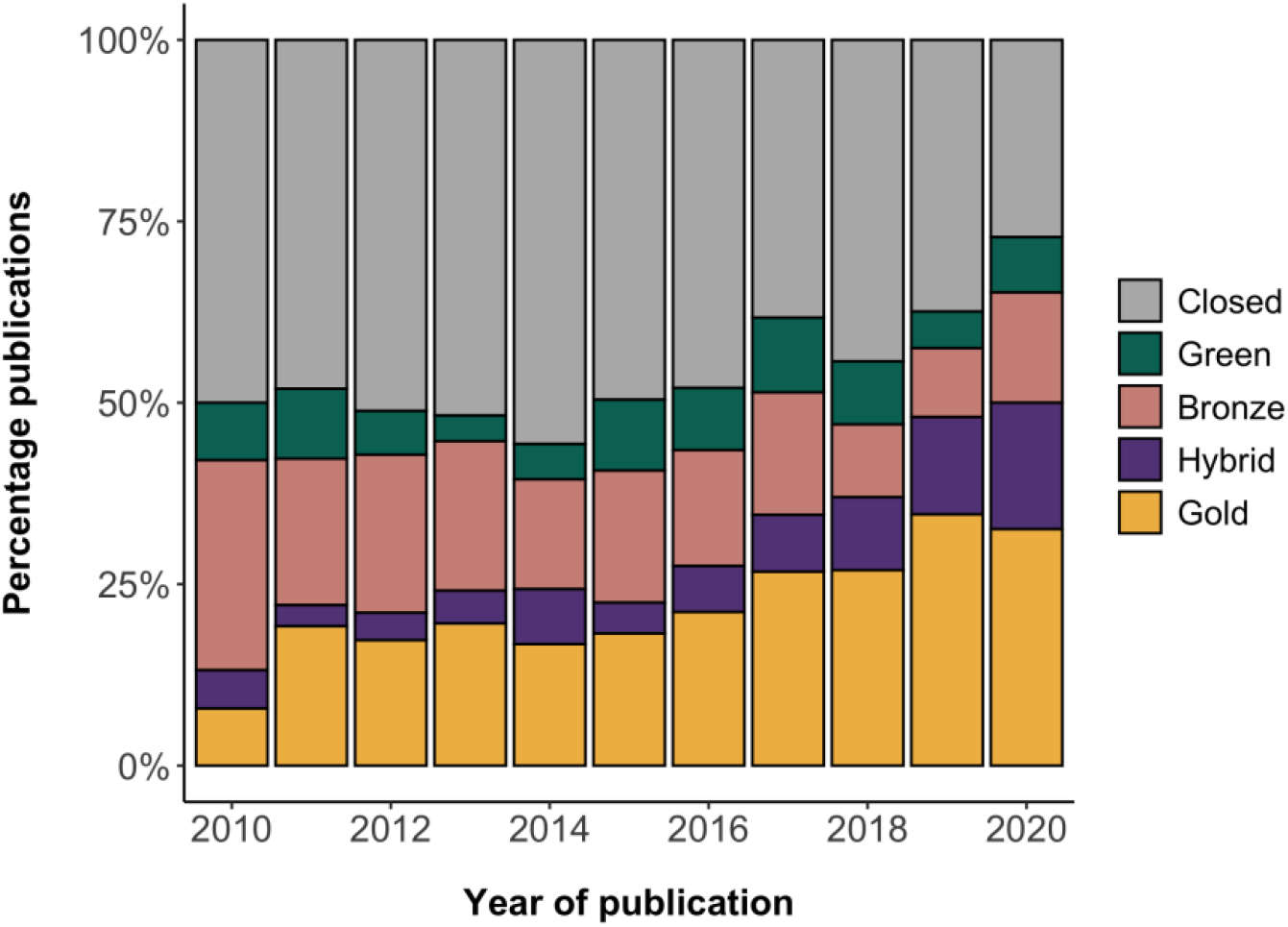
OA status of the clinical trial results publications in the cohort. Given the hierarchy used, green OA represents publications that were only openly accessible in a repository.

### 3.3. Self-archiving permissions of publications

Focusing on closed-access publications, 85% (n = 736/871, 95% CI 82% to 87%) had sufficient information to derive a self-archiving permission. The remaining closed-access publications for which a permission could not be determined include 35 (4%, 95% CI 3% to 6%) without a ‘best permission’ in the API response, 97 (11%, 95% CI 9% to 13%) with no information on the embargo period, and 3 (0.3%, 95% CI 0% to 1%) with insufficient information on the version and location of self-archiving (dark blue in **Figure 2**).

**Figure 2.**
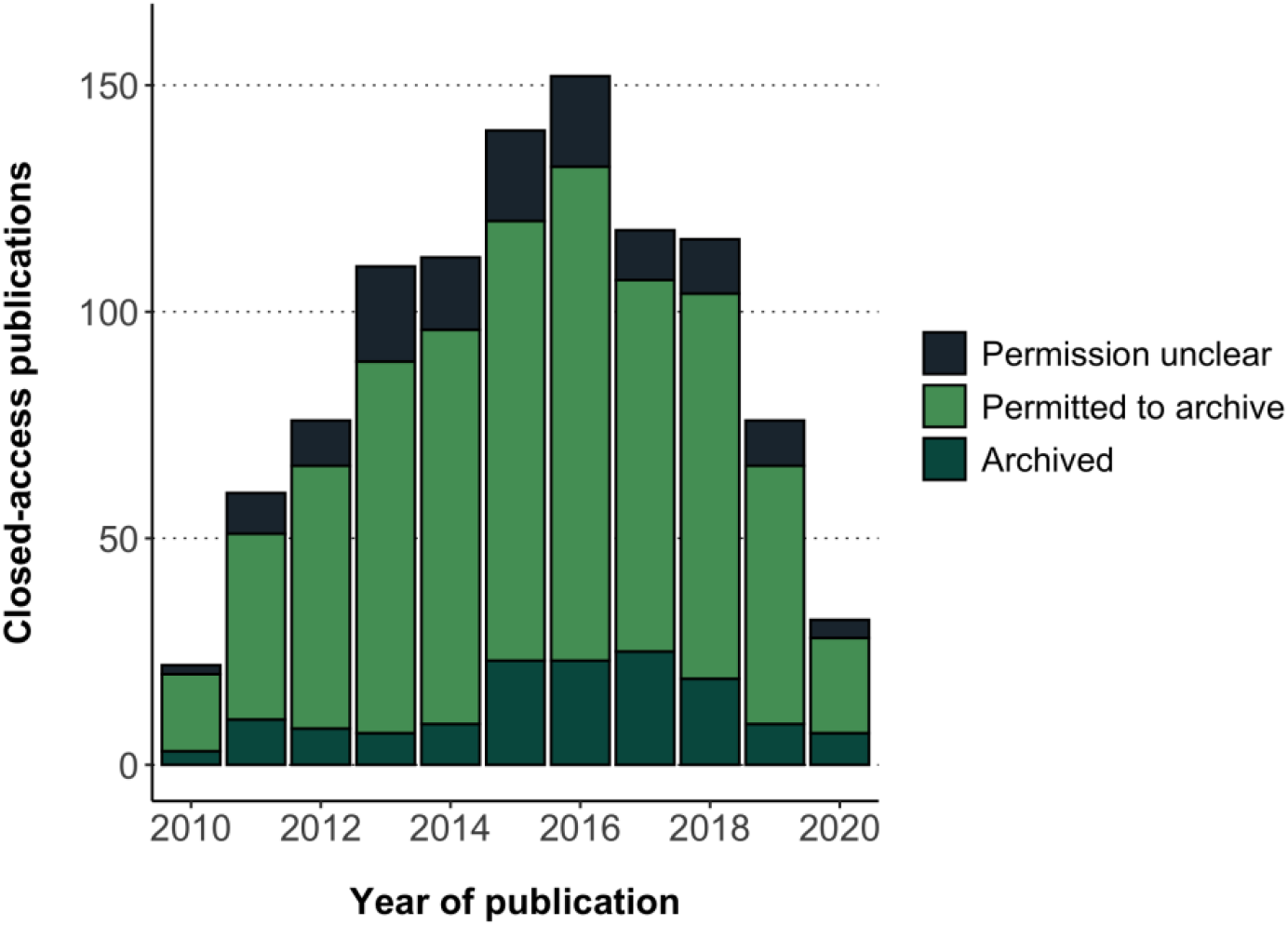
Realized potential of green OA for otherwise closed-access clinical trial results publications in the cohort.

Based on the criteria used in this study (**Table 1**), all 736 closed-access publications for which a self-archiving permission could be determined in Shareyourpaper had a permission to archive either the accepted or published version in an institutional repository (light green in **Figure 2**). More specifically, this included permissions to archive the accepted version only (n = 672), the published version only (n = 40), and both the accepted and published version (n = 24). Embargoes associated with these self-archiving permissions ranged between 0 – 24 months, with most publications having an embargo of 12 months (n = 636) (**Figure B3**). Taken together, 85% (n = 736/871, 95% CI 82% to 87%) of the closed-access publications in this cohort could be made accessible in an institutional repository, in large part based on permissions issued by journals or publishers to self-archive the accepted version after an embargo of 12 months (RQ2).

The realized potential of green OA for otherwise closed-access articles was estimated based on the number of green OA publications (n = 143) and the number of closed-access publications for which a self-archiving permission was found (n = 736). Thus, 143 of 879 (736 + 143) of otherwise closed-access publications with a self-archiving permission were made openly accessible in a repository (dark green in **Figure 2**). This corresponds to 16% (95% CI 14% to 19%) realized green OA in this cohort across all publication years.

## 4. Discussion

### 4.1. Overall discussion

This study leveraged open tools to assess the potential of green OA to broaden access to research and generate actionable information on self-archiving at scale. This was demonstrated in a cohort of results publications from clinical trials conducted at German university medical centers. Of the 1,897 clinical trial publications published between 2010 and 2020, 46% (n = 871/1,897) were not openly accessible via a journal or repository. Of these, 85% (n = 736/871) had a permission to self-archive the postprint in an institutional repository. Thus, many of the closed-access publications in this cohort could be made openly accessible in a location that supports long-term preservation, in line with WHO guidelines [25].

These findings corroborate the largely unrealized potential of green OA found in previous studies [14,18,36]. One study found that 39.2% of a cohort of global health research articles published in journals that allowed self-archiving had been made available via this route [36]. This is higher than the 16% realized green OA found in the present study, and may partly reflect the inclusion of a broader range of locations considered as green OA that are not harvested by Unpaywall (e.g., academic social networks). In any case, the gap between opportunity and practice is surprising, given both the practical and ethical relevance of clinical trials and the demonstrated higher impact of self-archived papers compared to paywalled papers based on citations [36,37]. Besides some of the known barriers to self-archiving, such as lack of awareness of self-archiving [15], the unrealized potential of green OA in this cohort likely also reflects the policy context and particularities in the research system in Germany. While many research and funding institutions in Germany have committed to OA, few institutional OA policies mandate self-archiving. This contrasts with other countries such as the UK, where high levels of green OA levels have been linked to OA mandates within the Research Excellence Framework [38,39], and the US where the White House Office of Science and Technology Policy recently updated policy guidance to make federally funded research freely available without delay [40]. Moreover, in Germany researchers based at universities have a high degree of autonomy, which makes it difficult to enforce compliance [3]. The outcome of as yet unresolved legal disputes relating to the right of secondary publication introduced in German copyright law will likely shape future efforts to promote self-archiving. A pilot to promote the use of such a clause in the Netherlands (“Taverne Amendment”) led to almost 3,000 publications being deposited through institutional repositories [13].

Comparisons with other studies are complicated due to different approaches, operationalizations of green OA, and focus on different disciplines. However, the overall OA share of 54% in this cohort of clinical trial publications is higher than the 43% OA share for universities in Germany between 2010 – 2018 obtained in a recent study [3]. In terms of the share of exclusive green OA, the finding that 8% of publications were green OA is in line with a previous analysis of the overall literature using Unpaywall, which used the same definition of green OA (available in the repository and not in the journal) [2]. Higher estimates of exclusive green OA in other studies (e.g., of 12% [9] and 27.2% [36]) likely reflect the inclusion of non-repository deposit locations (e.g., personal websites).

This study is novel in its use of Shareyourpaper to obtain self-archiving permissions at the level of individual articles. Shareyourpaper aims to derive the most advantageous permission to legally self-archive a paper, based on all the relevant policies that may apply for that paper. This includes funder mandates and institutional OA policies. There is increasing interest in monitoring OA at the institutional level and evaluating the impact of interventions on OA uptake [38]. Shareyourpaper seems like a meaningful tool to support such efforts, also given that the underlying data is open and fully machine-readable. Moreover, Shareyourpaper narrows the gap between opportunity and practice by automating the deposit workflow (including metadata entry, permissions and version checking). This makes it possible to act on self-archiving permissions obtained at levels beyond that of an individual institution (e.g., clinical trials across multiple institutions), and thus empowers efforts to increase self-archiving at scale.

The dissemination and open availability of clinical trial results is essential to support evidence-based decision-making by providers and patients alike, and fulfill ethical obligations to study participants. A previous study focusing on clinical trials in this cohort completed between 2014 – 2017 found that 30% had not reported results five years after trial completion [32]. Non-reporting of clinical trial results distorts our understanding of the medical evidence base, hampers evidence synthesis, and undermines medical decision-making [41]. Restricting access to clinical trial results behind paywalls risks further exacerbating these issues. Based on the OA definition used, the findings in this study suggest that compliance with the WHO guideline to make results publications openly accessible *where possible* is low. Most of the closed-access publications in this cohort had a permission to self-archive the accepted version in an institutional repository 12 months after publication. Yet, in most cases this possibility was not exploited to increase discoverability. The approach described in this study is being used as part of an ongoing pilot intervention at the Charité to improve clinical trial transparency, including leveraging publisher self-archiving permissions to make trial results publications openly accessible [42]. In brief, we developed and disseminated trial-specific report cards with feedback on a trial’s transparency and recommendations for improvement. If a clinical trial is found to have a results publication that is not accessible in the journal but could be self-archived, the report card recommends researchers to self-archive the publication using Shareyourpaper or by contacting their institutional library.

### 4.2. Strengths and Limitations

One of the strengths of this study is the development of a fully automated approach based on open tools (Unpaywall and Shareyourpaper) to generate actionable information on self-archiving. This approach can empower efforts to increase science discoverability at scale. The underlying code used to query the APIs is openly available and can be adapted for further use, including the application of different criteria to derive self-archiving permissions. Furthermore, this approach was demonstrated in a validated cohort of clinical trial results publications at the level of German university medical centers. Assessing the realized potential of green OA at this level is meaningful for several reasons: 1) sharing the results of clinical trials is of ethical and practical relevance, 2) WHO guidelines state that clinical trial results publications should be openly accessible, where possible, and 3) an analysis at this level can inform interventions to increase self-archiving at a broad scale while also allowing the impact of institutional policies to be evaluated.

This approach also faces limitations. The findings depend on the information in Unpaywall and Shareyourpaper being accurate and up-to-date. Unpaywall has previously been shown to provide a conservative estimate of the actual percentage of OA in the literature [2]. In turn, a recent study found changes in Unpaywall OA classifications over time, which may reflect previous errors or a true change in OA status [43]. However, it is unclear whether this affected this study, also because OA classifications were defined in this study via UnpaywallR (**Table A1**). Taken together, some publications that were reported as closed may in fact have been openly accessible. Moreover, based on the OA definition used in this study, publications that were only free-to-read in a location other than a journal or repository (e.g., ResearchGate) were not considered as OA. Publications with no ‘best permission’ or no embargo information in Shareyourpaper may have been archivable, and other self-archiving routes beyond those examined in this study could be considered. Moreover, while Shareyourpaper takes funder mandates and institutional OA policies into account, they were not considered in this study. Thus, the potential of self-archiving to increase discoverability may be higher than shown here. Finally, the proportion of realized green OA reported in this study is an estimate, since: a) green OA publications were assumed to have had a permission for self-archiving, and b) neither the version nor the deposit location of green OA publications was checked, and may have included other forms of deposit beyond self-archiving of the postprint in an institutional repository.

### 4.3. Future research directions

Libraries may adapt the approach described in this study to support their efforts to populate institutional repositories. This could be achieved by promoting the use of Shareyourpaper among authors at their institution, or by embedding the described tools into existing self-archiving workflows. One strength of this approach lies in the ability to relatively quickly flag publications that can be made openly accessible via self-archiving. Providing an institutional record of publications exists, this could serve to identify the target group for institutional initiatives that aim to raise awareness of and promote self-archiving, thereby increasing their impact. This includes training workshops and consultation services, but also targeted interventions to promote deposits to an institutional repository (e.g., during Open Access week). Whether implementing such an intervention would entail additional resources would depend on its scale and the integration of such tools. A first step may be to pilot such an intervention with publications that have a permission to self-archive the Version of Record.

This approach may also inform interventions to provide access to closed-access publications at other levels beyond that of a given institution. For example, an intervention at the level of protocols published in toll-access journals could be particularly meaningful, given that being able to reproduce the methods described in these publications depends on having access to them. Shareyourpaper provides the date at which an embargo will elapse (if applicable) and supports deferred deposits for embargoed articles in Zenodo. Therefore, such interventions could in principle promote future self-archiving already at the time of publication. This would spare researchers the hassle of finding the version of their publication that can be shared long after publication, and could help embed self-archiving as an integral phase of the publication process. Finally, while arguably the priority is to make closed-access publications openly accessible, this approach could also inform efforts to increase self-archiving of OA publications to strengthen the long-term preservation of research outputs.

## Data Availability

All the code generated in this study is available in GitHub under an open license (https://github.com/delwen/oa-archiving-permissions). The data presented in this study are openly available in Zenodo at https://doi.org/10.5281/zenodo.7455305.

https://github.com/delwen/oa-archiving-permissions

## Supplementary Materials

See Appendix A-B.

## Author Contributions

Conceptualization, DLF; methodology, DLF; software, DLF; formal analysis, DLF; investigation, DLF; data curation, DLF; writing—original draft preparation, DLF; writing— review and editing, DLF; visualization, DLF.

## Funding

This research was funded by the Bundesministerium für Bildung und Forschung, grant number 01PW18012.

## Data Availability Statement

All the code generated in this study is available in GitHub under an open license (https://github.com/delwen/oa-archiving-permissions). The data presented in this study are openly available in Zenodo at https://doi.org/10.5281/zenodo.7455305 [35].

## Acknowledgments

This work was informed by valuable and constructive input from Maia Salholz-Hillel and Daniel Strech.

## Conflicts of Interest

The authors declare no conflict of interest. The funders had no role in the design of the study; in the collection, analyses, or interpretation of data; in the writing of the manuscript; or in the decision to publish the results.

## Appendix A: Supplementary Tables

**Table A1.** Classification of OA types.

| **OA type** | **Description** | **Combination of fields** |
| --- | --- | --- |
| Closed | The publication is not openly accessible | `is_oa = FALSE OR`<br>`oa_locations is NULL` |
| Gold | The publication is free to read in a fully OA journal | `is_oa = TRUE AND host_type`<br>`= "publisher" AND`<br>`journal_is_oa = TRUE` |
| Hybrid | The publication is free to read under an open license in a non-fully OA journal | `is_oa = TRUE AND host_type`<br>`= "publisher" AND`<br>`journal_is_oa = FALSE`<br>`AND license in oa_locations`<br>`starts with "cc-"` |
| Bronze | The publication is free to read without a clearly identifiable license in a non-fully OA journal | `is_oa = TRUE AND host_type`<br>`= "publisher" AND`<br>`journal_is_oa = FALSE` |
| Green | The publication is free to read in a repository (institutional, subject-specific, or other type of repository) | `is_oa = TRUE AND host_type`<br>`= "repository"` |

\* Description of the OA types used in this study, along with the combination of fields to derive them in UnpaywallR.

## Appendix B: Supplementary Figures

**Figure B1.**
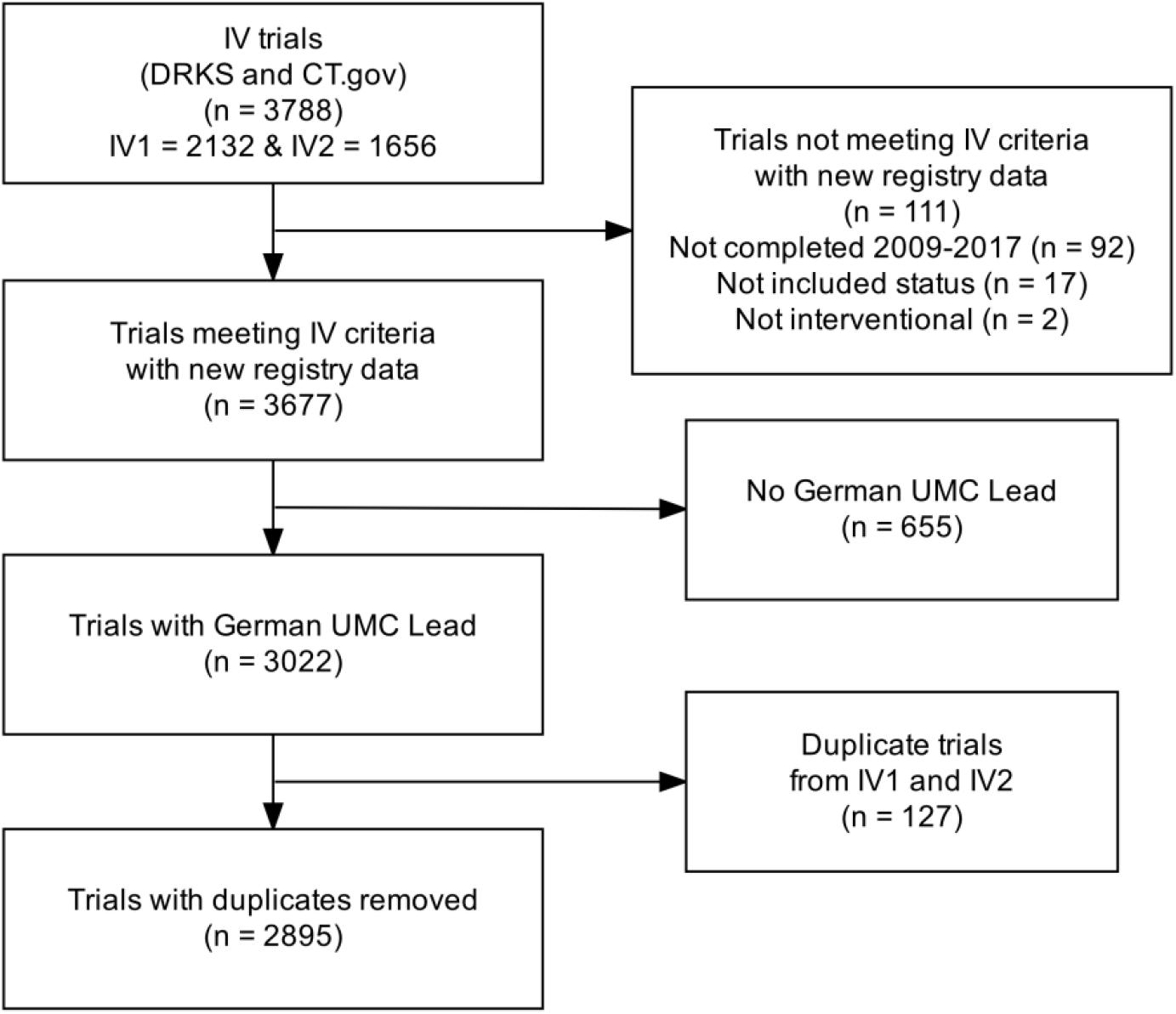
Flow diagram of the trial screening. Abbreviations: CT.gov: ClinicalTrials.gov; DRKS: German Clinical Trials Register; IV: IntoValue; UMC: university medical center.

**Figure B2.**
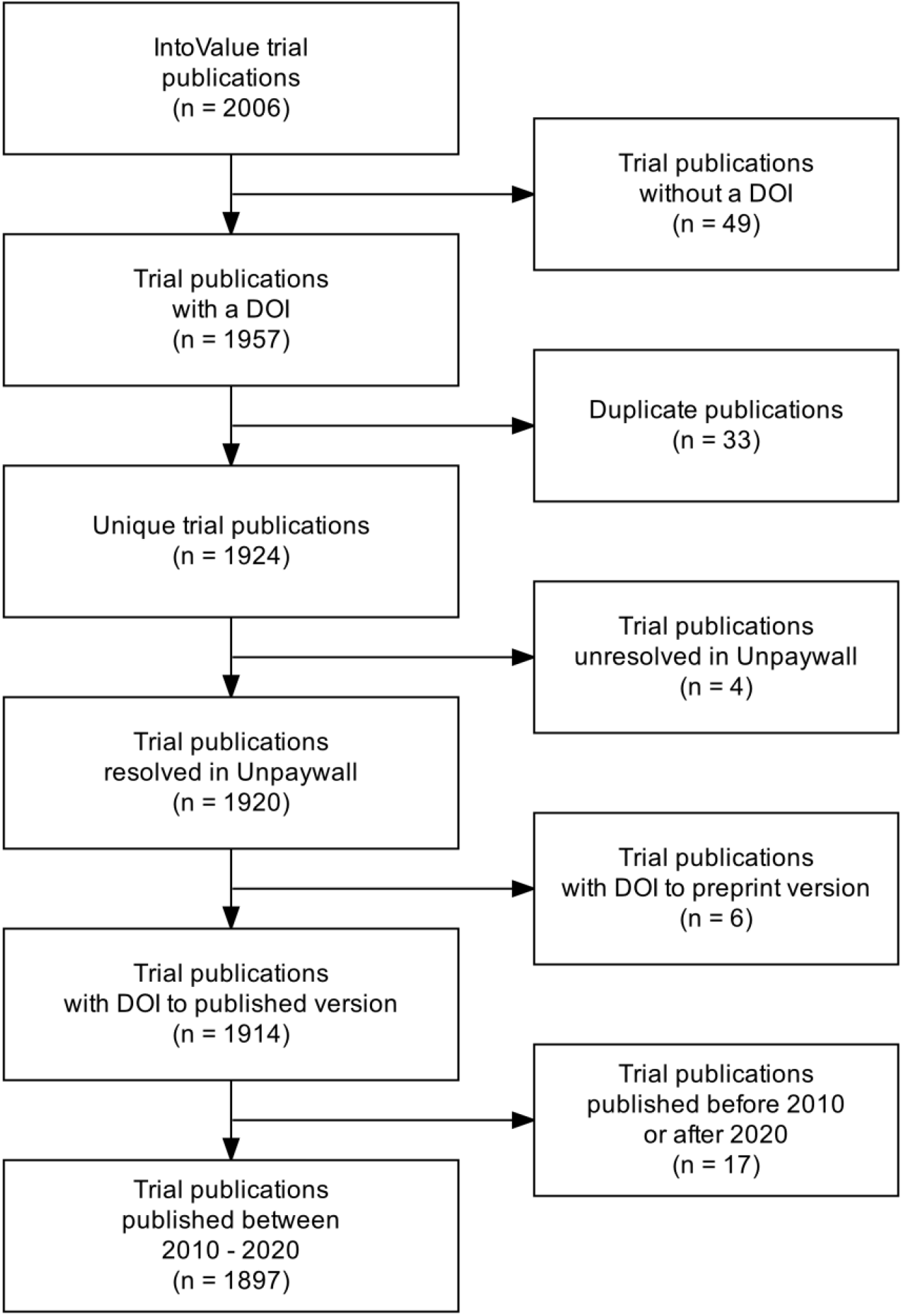
Flow diagram of the publication screening. Abbreviations: DOI: digital object identifier.

**Figure B3.**
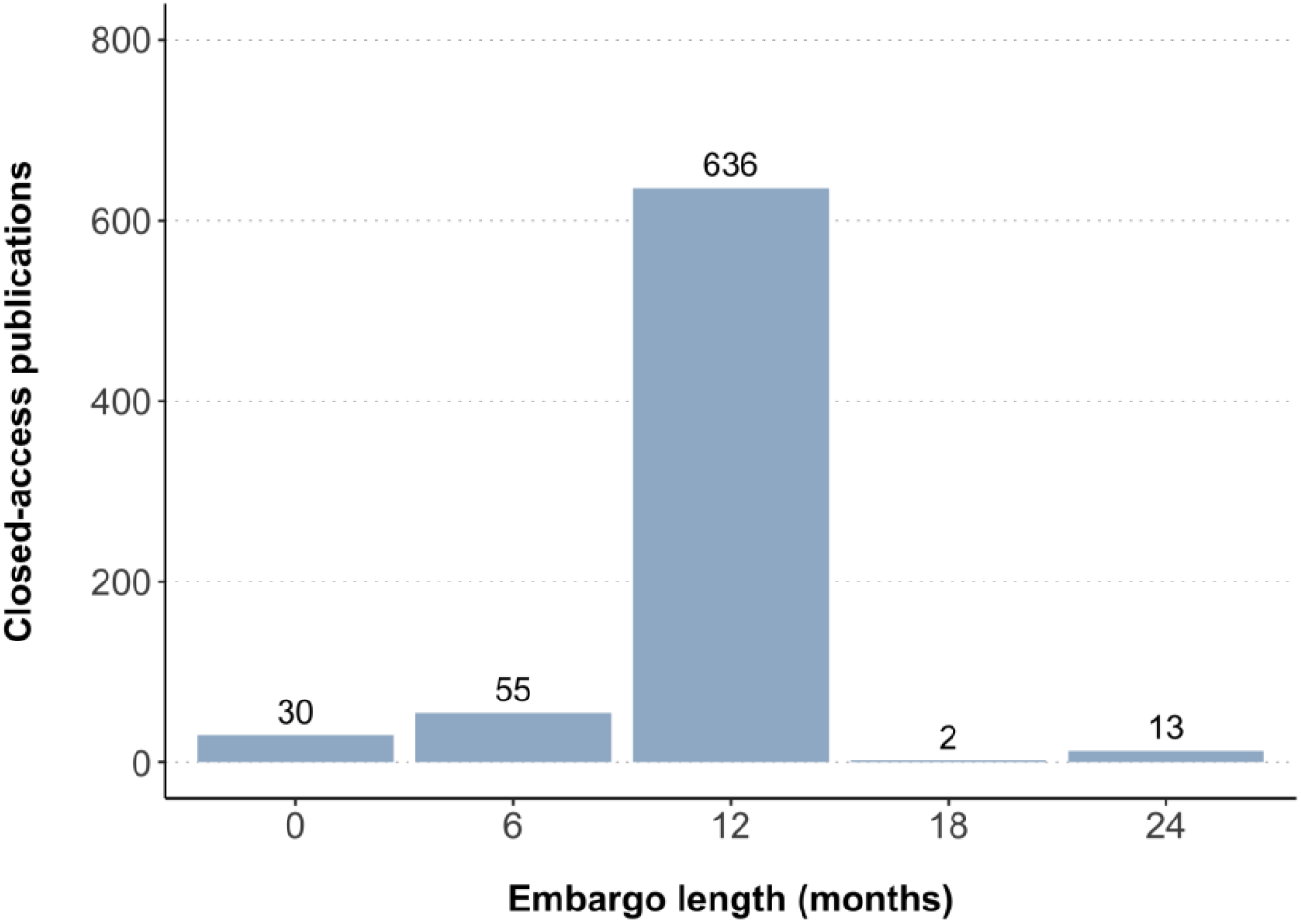
Embargo periods associated with self-archiving permissions found for closed-access publications.

## Notes

### Competing Interest Statement

The authors have declared no competing interest.

